# Automating the triage of rheumatology outpatient referrals: a comparative evaluation of 23 large language models under simple and advanced prompting

**DOI:** 10.64898/2026.08.05.26359488

**Authors:** Lynden Roberts

## Abstract

**Objective:** Triage of rheumatology outpatient referrals is a high-volume administrative task that consumes senior specialist time without advancing patient care. The human triage system is only moderately accurate and reproducible. We assessed whether contemporary large language models (LLMs) are able to perform well enough to support automating this task in practice. In addition, the effects of different prompting techniques on triage accuracy and cost was assessed to help identify to optimal approach.

**Methods:** Twenty referral scenarios spanning the urgency spectrum, based on real referrals were created by a certified Australian rheumatologist. Four rheumatologists triaged all cases independently and blinded, to produce a consensus reference standard. Twenty-three LLMs each triaged every referral into one of five urgency categories, three times (1380 outputs per condition). The experiment was run with a simple prompt and repeated with a advanced prompt supplying explicit triage expectations and worked examples.

**Results:** All 2760 attempts returned valid categories. Under the simple prompt, performance separated into distinct tiers, larger models were more accurate (Spearman rho=0.42; P=.047) and accuracy tracked cost. Advanced prompting minimised between-model variance in accuracy 5.3-fold (0.014 to 0.003; Levene P=.01), abolished the size-accuracy association (rho=-0.05; P=.83) and removed the accuracy-cost relationship. Leading models matched expert consensus on most cases, within or above the range reported for human triage. Under-triage errors persisted with some LLMs.

**Conclusion:** Contemporary LLMs categorise rheumatology referral urgency as well or better than published human triage systems. Advanced LLM prompting methods substitute for the reasoning capability of larger models, suggesting that LLM performance on this task may not require the most expensive models. The tools to automate this administrative task appear to already exist. Strong candidate LLMs that might serve a production ready solution have been identified.

## Introduction

Safe access to rheumatology outpatient care depends on triage: the sorting of incoming referrals by clinical urgency so that finite specialist capacity is directed first to those at greatest risk of harm from delay. In most services this is done by a consultant rheumatologist or senior registrar reading free-text referral letters. It is a high volume, cognitively demanding, and it a time-consuming task using clinician resources that does not itself advance a patient’s care or generate any therapeutic outcome [1, 2]. Every hour spent triaging is an hour lost to direct clinical work. As referral volumes rise against a rheumatology workforce that is widely projected to fall short of demand [3], this opportunity cost has become a pressing operational problem for the specialty rather than a peripheral administrative irritation.

The rheumatology casemix exemplifies the risks and costs of referral triage systems. The spectrum reaching a clinic ranges from organ- or sight-threatening emergencies such as giant cell arteritis, systemic vasculitis and lupus with organ involvement, through to degenerative and non-inflammatory conditions that can safely wait months [4]. Getting urgency wrong carries real consequences: delays in the assessment of early inflammatory arthritis are associated with worse long-term joint and functional outcomes [5].

Manual triage is also an imperfect standard against which to judge an automated alternative. In one rheumatology series the urgency grade assigned from the referral changed after specialist consultation in almost half of cases, and roughly one in six truly urgent patients were under-triaged at the referral stage, creating avoidable clinical risk [6]. A structured rheumatology referral tool detected urgent presentations with a sensitivity and specificity of 80% and 79% respectively, leading to problematically high rates of misclassification [7]. Reproducibility is similarly mediocre: independent clinicians prioritising identical referrals typically agree only moderately (weighted kappa about 0.6), disagree on roughly a third of cases, and do not improve appreciably with training [8, 9], while specialists vary widely when triaging identical referrals across settings [10]. The pertinent question for an automated aid is therefore not whether it is perfect, but whether it matches or exceeds a human standard that is itself only moderately accurate and reproducible - particularly given automation advantages of speed, turnaround, consistency and cost.

Published approaches to automated triage have relied on conventional natural language processing and machine learning applied to referral letters, delivering modest, narrowly scoped gains insufficient for rollout at scale [4, 11]. Large language models (LLMs) appear better suited: they encode broad clinical knowledge, pass licensing examinations, and can support diagnostic and triage decisions across specialties [12–14]. Early referral-triage evaluations in other specialties report useful but imperfect accuracy [15], and within rheumatology LLMs have approached specialist performance on diagnostic tasks [16]. A recurring theme is that how a model is prompted - chain-of-thought reasoning, worked examples, guided reflection - can matter as much as which model is used [17–19]. No published study has yet interrogated how prompt design interacts with model capability and cost across a broad contemporary panel on specialist urgency triage.

This gap has direct economic consequences for deployment at scale. If triage accuracy tracks model size and price, achieving affordability may require acceptance of poorer performance; if a well-specified prompt can raise smaller, cheaper models to competitive accuracy, the economics of a deployable triage assistant may prove viable. We therefore benchmarked 23 contemporary LLMs on the urgency triage of typical rheumatology outpatient referrals against a blinded four-rheumatologist consensus, comparing their performance, its relationship to serving cost, and the effect of two prompting strategies.

## Methods

### Study design and setting

A comparative evaluation study at a specialist rheumatology clinic within an Australian academic healthcare network was conducted. The task under evaluation was the assignment of an urgency triage category to a general practitioner (GP) referral, mirroring the administrative decision made daily in the service. Every model triaged every case, each case was attempted three times, and the entire experiment was repeated under two prompt conditions. Thus, 23 models x 20 cases x 3 attempts = 1380 model outputs per prompt condition, and 2760 outputs in total.

### Case selection and development

Twenty referrals representative of the range of presentations typically received by the clinic from Australian GPs were constructed by certified rheumatologists, who have personally triaged many thousands of such referrals. Each scenario was written to be consistent with real referral letters in content, clinical detail and level of completeness. The set was purpose-built to span the full urgency spectrum and the common diagnostic categories seen in practice: from organ- or life-threatening inflammatory and autoimmune disease requiring near-immediate review, through active but stable inflammatory conditions, to degenerative and non-inflammatory presentations appropriate for routine scheduling.

### Triage categories and definitions

Each referral was assigned to one of five mutually exclusive urgency categories, defined by the maximum safe waiting time: Emergency (seen within 1 day), Hyper-Urgent (within 7 days), Urgent (within 1 month), Semi-Urgent (within 3 months) and Routine (within 12 months). The governing principle, applied by both human experts and models, was to select the category that achieved the safest longest wait, without risking death or permanent damage.

### Gold-standard consensus

The reference standard for each case was established by four rheumatologists who each triaged all 20 referrals independently, blinded to one another’s decisions and to model outputs. A single consensus urgency category per case was then agreed and used as the gold standard against which all model outputs were scored. For a small number of cases the expert reference concluded that either of two adjacent categories represented correct triage.

### Language models and serving platform

Twenty-three contemporary LLMs were evaluated, and the identical model set was used in both prompt conditions. Models were accessed through a single application programming interface aggregator (OpenRouter), which provides unified access to models from multiple providers. The evaluated models were: anthropic/claude-fable-5, anthropic/claude-haiku-4.5, anthropic/claude-opus-4.8, anthropic/claude-sonnet-5, deepseek/deepseek-v4-pro, google/gemini-2.5-flash, google/gemini-2.5-pro, google/gemini-3.1-pro-preview, google/gemini-3.5-flash, meta-llama/llama-4-maverick, minimax/minimax-m3, mistralai/mistral-medium-3-5, moonshotai/kimi-k2.6, nvidia/nemotron-3-ultra-550b-a55b, openai/gpt-5.4-nano, openai/gpt-5.5, openai/o1, openai/o3, openai/o4-mini-high, qwen/qwen3.6-flash, qwen/qwen3.7-max, x-ai/grok-4.3 and z-ai/glm-5.2. Hence, the panel spanned a wide range of model families, sizes, release generations and serving prices, including both large frontier reasoning models and smaller, lower-cost models.

Experiments were run via a purpose-built bulk-testing platform (internally developed web application) designed to test prompts, cases and models at scale. The platform issued each model-case-attempt request, captured the model’s chosen category and free-text rationale, and logged latency, prompt and completion token counts, total tokens and serving cost for every attempt. Serving cost is reported as the price charged by the aggregator for model inference, in US dollars (USD).

### Prompt conditions

Two prompts were evaluated (full text in Table 1). The simple prompt provided minimal task instructions. The advanced prompt added explicit direction reflecting Australian triage expectations, instructed chain-of-thought reasoning, directed the model to focus on biological disease by disregarding pain severity and psychosocial impact, and supplied worked examples of conditions belonging to each category.

### Repeat runs

To test output stability, each model triaged each case three times, separately and independent of prior result knowledge, within each prompt condition. Repeated sampling allowed each model decision to be summarised as a confidence profile rather than a single point estimate, distinguishing decisions reached unanimously across attempts from those reached less consistently.

### Outcomes and scoring

The primary outcome was triage accuracy relative to expert consensus. For each model and case, the three attempts were collapsed into a repeat-run confidence profile with five mutually exclusive outcomes: high-confidence correct (matched consensus on all three attempts, 3/3); lower-confidence correct (matched on 2 of 3); miss by over-triage (more urgent category than the expert); miss by under-triage (less urgent category than the expert); and unclassified or error. A case was counted as correct when the model’s category matched consensus on the majority of runs, and per-model accuracy was the proportion of correct cases. The distinction between over- and under-triage was made there is asymmetric clinical risk: over-triage is safer, whereas under-triage exposes a patient to potentially harmful delay.

Models were ranked by total correct (3/3 + 2/3), then by high-confidence correct, then by fewer over-triage misses, fewer under-triage misses, and fewer unclassified or error outcomes; remaining ties were broken by lower total run cost.

### Statistical analysis

Results were summarised descriptively and visually. For each prompt condition we examined the distribution of confidence-profile outcomes across the 23 models (Figs. 1, 2) and the relationship between per-model accuracy and per-model serving cost (Figs. 3, 4). Associations between model size, cost and accuracy were assessed with Spearman rank correlation. Equality of between-model variance in accuracy across the two prompt conditions was tested with the Levene test, with the Bartlett test as a sensitivity analysis. Two-sided P values below .05 were considered statistically significant.

**Fig. 1.**
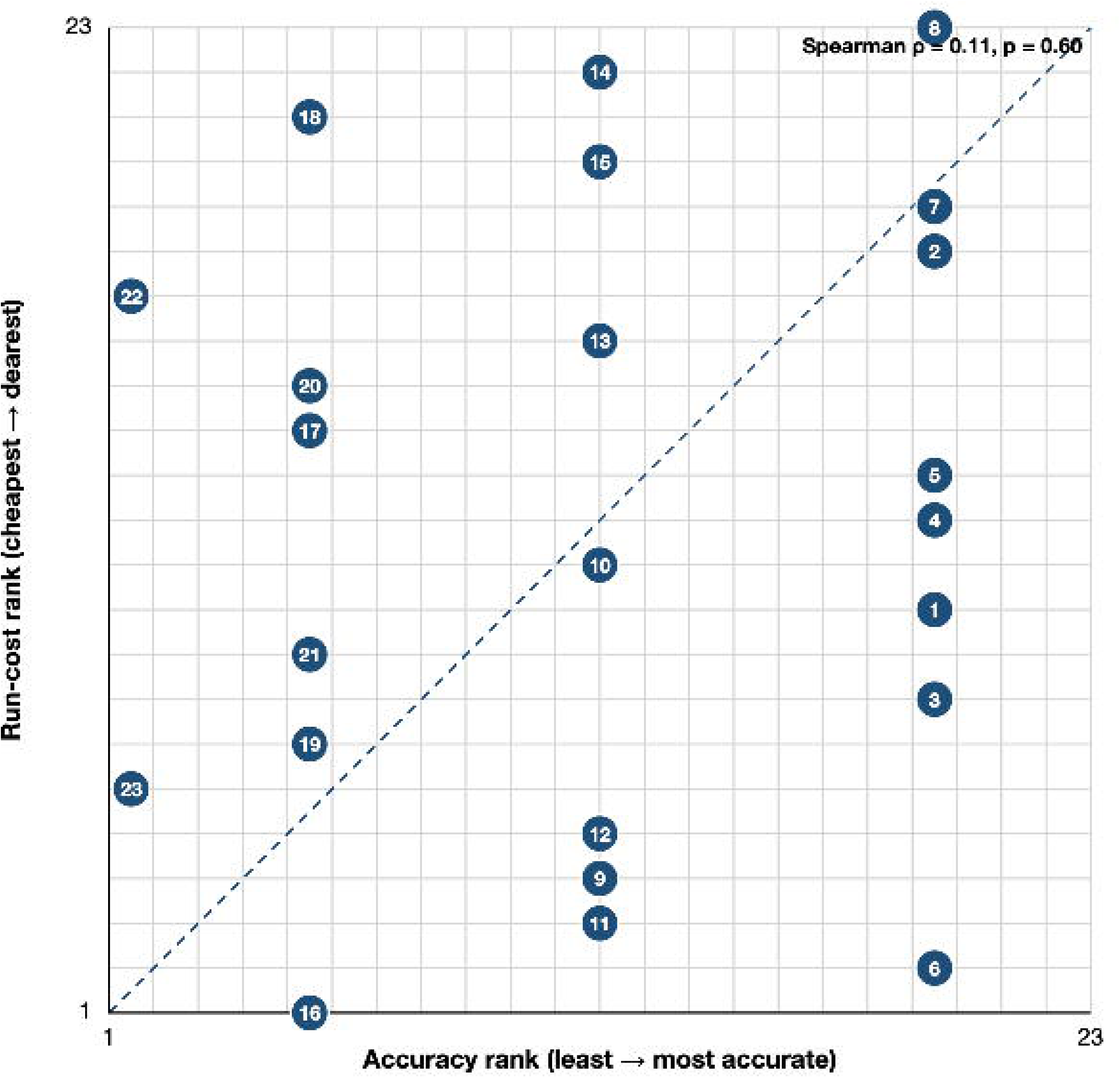
Triage performance of 23 large language models under the simple prompt. Each row shows one model. Stacked bars show the repeat-run confidence profile across three attempts per referral: high-confidence correct (matched expert consensus on 3/3 attempts), lower-confidence correct (2/3), miss by over-triage (more urgent category than consensus), miss by under-triage (less urgent category than consensus), and unclassified or error. Right-hand columns show total serving cost in US dollars (USD), days since the model’s public release calculated to 1 July 2026, and model size in parameters (best available estimate; inequality signs denote undisclosed commercial models). Models are ranked by total correct.

**Fig. 2.**
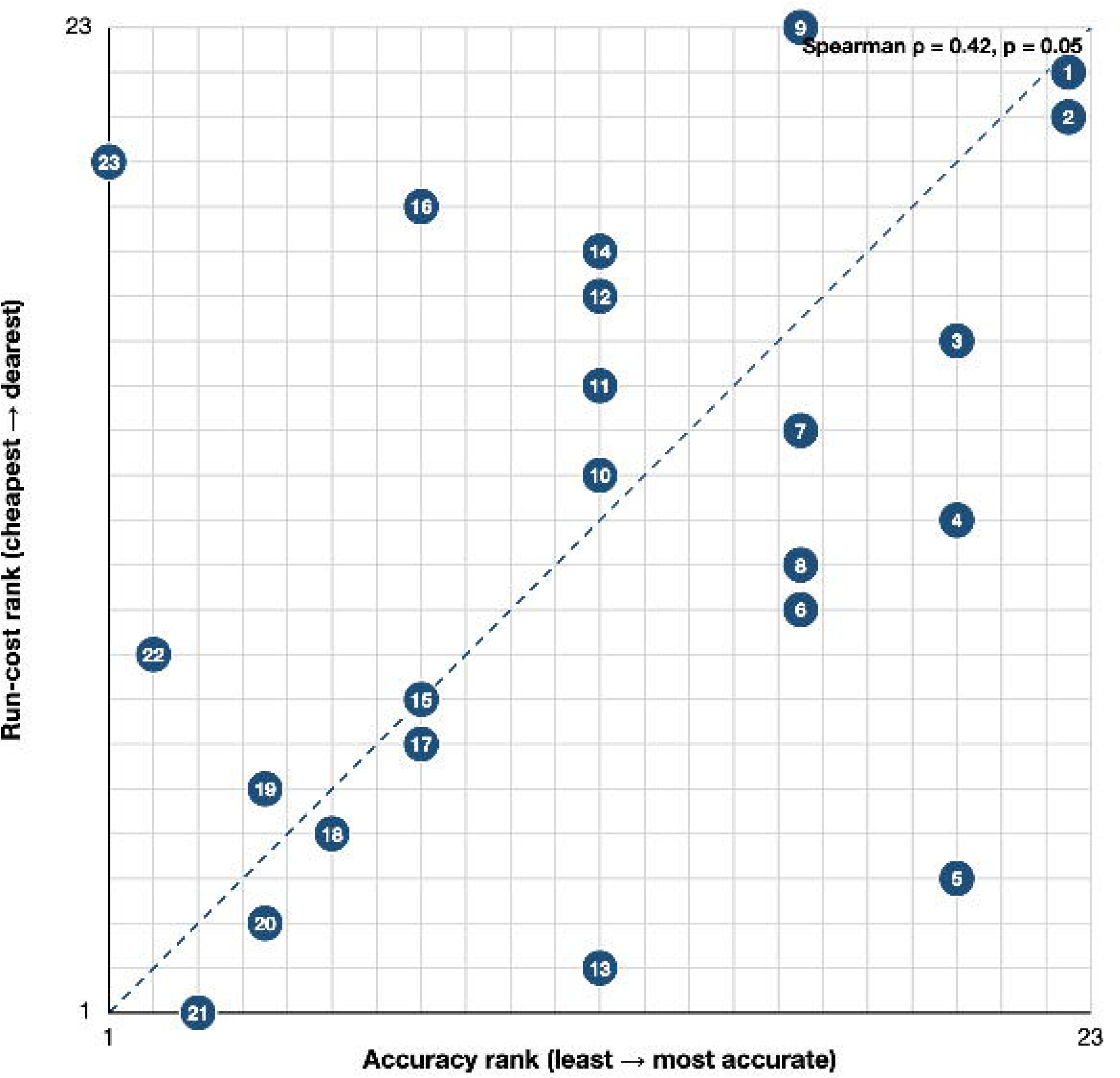
Triage performance of the same 23 large language models under the advanced prompt. Categories, right-hand columns and ranking are as in Fig. 1. Days since release are calculated to 1 July 2026 and cost is reported in US dollars (USD). Between-model variance in accuracy is markedly compressed relative to Fig. 1.

**Fig. 3.**
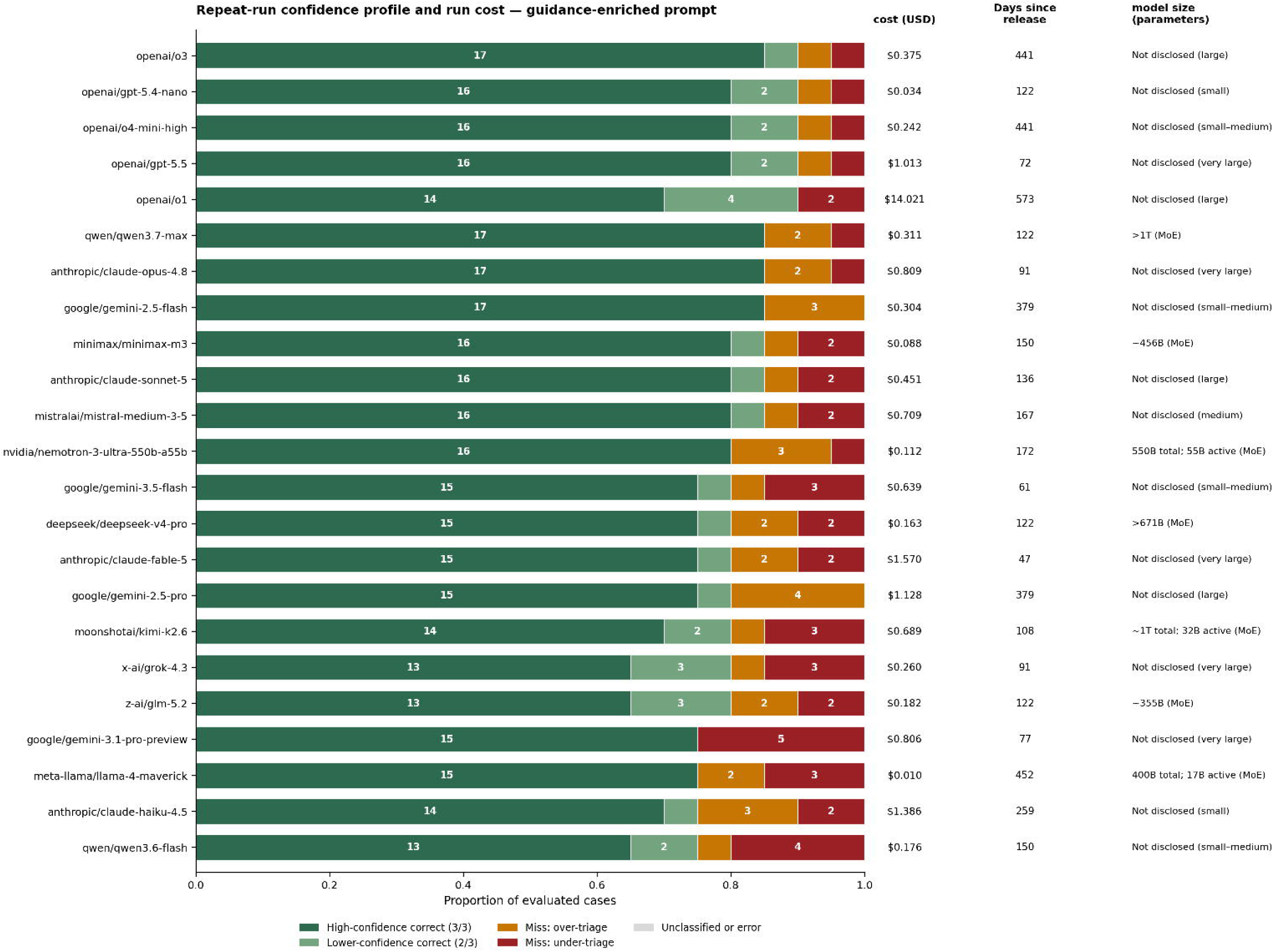
Association between per-model triage accuracy and per-model serving cost under the simple prompt. Each point represents one model. Accuracy increased with cost (Spearman rho=0.42; P=.05).

**Fig. 4.**
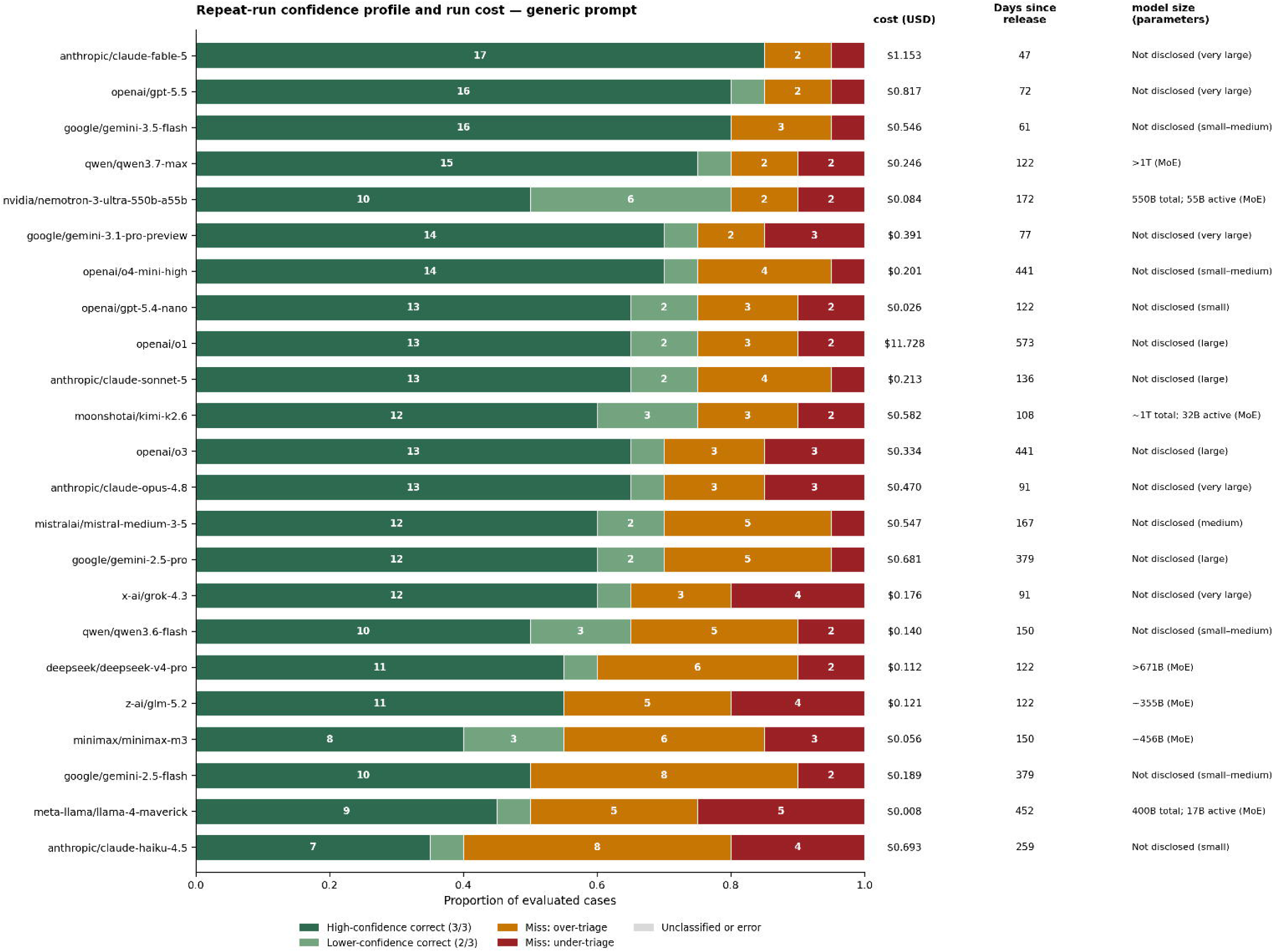
Association between per-model triage accuracy and per-model serving cost under the advanced prompt. Each point represents one model. No systematic accuracy advantage was seen for higher-cost models (Spearman rho=0.11; P=.60).

### Ethical considerations

The study was conducted under an ethics waiver granted by the Monash Health Human Research Ethics Committee, Monash Health, Melbourne, Victoria, Australia, and followed the principles of the Declaration of Helsinki. No patient-identifiable information was used in the study or provided to any external model provider.

## Results

### Overview

The 23 models returned valid triage categories in all 1380 model-case-attempts in each prompt condition. Performance varied widely across the LLMs, and both the level of performance and its relationship to model cost differed markedly between the two prompt conditions.

### Model capability spanned a wide range under the simple prompt

Under the simple prompt, the LLMs separated into clearly distinct tiers of performance (Fig. 1). Larger and more recent models achieved the highest proportions of correct cases, with a substantial share of their correct answers reached unanimously across all three attempts. Smaller and older models performed considerably less well, with fewer total correct cases, a smaller unanimous fraction and a greater burden of misclassification. The spread between the strongest and weakest models was large, indicating that, when given simple instruction, the ability to translate a free-text referral into a safe urgency category was variable across contemporary LLMs. Across the LLMs, larger models achieved higher accuracy under this prompt (Spearman rho=0.42; P=.047).

### Advanced prompting levelled performance across the panel

Repeating the experiment with the advanced prompt narrowed the gap between models substantially (Fig. 2). Between-model variance in accuracy fell from 0.014 to 0.003, a 5.3-fold reduction (Levene test P=.01; Bartlett test P<.001). The smaller and older models that had lagged under the simple prompt improved considerably once the prompt supplied explicit triage expectations and worked category examples, moving closer to the leading models in total correct cases and in the proportion of high-confidence correct decisions. The strongest models remained strong, but their advantage was negated. The positive association between model size and accuracy seen under the simple prompt disappeared (Spearman rho=-0.05; P=.83).

### Accuracy was associated with cost under the simple prompt

Plotting per-model accuracy against per-model serving cost under the simple prompt revealed a positive association (Spearman rho=0.42; P=.05). More expensive models tended to achieve higher accuracy (Fig. 3). With minimal prompting instruction, therefore, paying more for a larger, more capable model bought better triage performance, and cost served as a partial proxy for accuracy on this task.

### The accuracy-cost association disappeared under advanced prompting

The same relationship was absent under the advanced prompt (Fig. 4). With explicit expectations and worked examples in the prompt, higher-cost models no longer showed a systematic accuracy advantage over lower-cost models (Spearman rho=0.11; P=.60). This dissociation is a key finding: advanced prompting techniques not only raised the performance of weaker models but severed the link between how much a model cost and how accurately it triaged.

### Errors were distributed between over- and under-triage

Across both conditions, residual misclassification was not confined to the safer direction. The confidence profiles (Figs. 1, 2) show that misses comprised both over-triage, in which a model assigned a more urgent category than the expert consensus, and under-triage, in which it assigned a less urgent category. Because under-triage is the direction that exposes a patient to potentially harmful delay, the persistence of under-triage errors - even among otherwise high-performing models and even under the advanced prompt - is the most safety-relevant feature of the results and directly shapes the requirements for any deployed system.

## Discussion

### Principal findings

We benchmarked 23 contemporary LLMs on the urgency triage of rheumatology outpatient referrals against a blinded four-rheumatologist consensus under two prompts. Four findings stand out. First, performance varied widely: contemporary LLMs are not interchangeable on specialist triage. Second, a advanced prompt with explicit expectations and worked examples produced a levelling effect, improving weaker, smaller and older models and compressing the performance gap 5.3-fold. Third, accuracy tracked serving cost under the simple prompt but not under the advanced prompt. Fourth, the leading models matched the consensus on most cases - unanimously across repeated runs for many - placing them within or above the range reported for human referral triage [6–10].

An interpretation is that under minimal instruction the model must supply, from its own capacity, both the clinical reasoning and the implicit triage policy needed to convert a referral into a safe category. Larger, more recent, more expensive models hold more of that latent capacity, so they perform better and cost more. When the prompt supplies the missing policy and reasoning scaffold, it substitutes for the capacity smaller models lack: the weaker models catch up and cost ceases to predict accuracy. Demonstrating this dynamic on a genuinely complex clinical-reasoning task is novel and practically consequential.

### Comparison with prior work

Our results extend evidence that prompt design can be as influential as model choice in clinical LLM applications. Structured prompting aligned model recommendations with an expert cardiology Heart Team far better than zero-shot prompting [18], and task-tailored, few-shot prompts raise performance on complex clinical tasks [19–21]. Our contribution is to show, across an unusually broad and contemporary LLM panel on a single specialist task, that advanced prompting not only raises average performance but specifically reduces the between-model differences and abolishes the accuracy-cost gradient - a pattern with direct procurement implications for services.

The findings also align with prior referral-triage work. Earlier studies showed LLMs could triage referrals with some accuracy, though performance remained uneven across presentations [15, 22]; machine-learning pipelines applied to GP letters could prioritise high-risk patients but were less effective when referrals were complex or ambiguous [4, 11]. Broader benchmarking finds capable but imperfect performance with both error directions and a consistent call for human oversight [14, 23]. Within rheumatology specifically, LLMs have already approached specialist performance on diagnostic reasoning [16]; the present study extends that finding from diagnosis to the operational decision that actually governs access to care.

### Why this matters to rheumatology services

Triage is, from the patient’s perspective, invisible work. It consumes senior rheumatologist time, generates no diagnosis, no treatment and no outcome, and yet must be done accurately because the cost of error falls on patients with time-critical inflammatory and autoimmune disease [1, 2, 5]. It is precisely the sort of task that should be automated first: high volume, rule-governed, repetitive, and non-therapeutic. The barrier has never been desirability but evidence that automation is safe enough. The present data address that barrier directly. The strongest models agreed with a four-rheumatologist consensus on most cases and did so reproducibly across repeated sampling, which places them at or above the band reported for human referral triage - a standard where urgency grades are frequently revised after assessment, a significant minority of urgent patients are initially under-triaged [6, 7], and independent clinicians disagree on around a third of cases with little benefit from training [8–10]. Unlike a clinician, a model can be resampled, audited, and does not tire, or suffer the cognitive biases that affect clinical judgement [16, 24]. Models carry their own risks of encoded bias, which must be audited rather than assumed absent [25].

The second finding changes the economics. Because a well-specified, example-based prompt lets smaller, cheaper models perform competitively, an affordable and scalable triage assistant no longer requires access to the most expensive frontier model. For a rheumatology service, this converts automation from a capital-intensive proposition into a realistic near-term one, and returns senior clinician hours to direct patient care at a time when workforce shortfalls are projected to worsen [3]. Our factorial platform offers a template for the local evaluation of prompts, cases and models that responsible deployment requires, including post-deployment auditing.

However, two caveats are worth considering here. Our reference standard is itself human judgement embedding one service’s norms, so ‘human-level’ means agreement with expert rheumatologists, not demonstrated superiority. And residual under-triage persisted even in the best condition. Any deployed system should therefore be configured to fail safe - biased towards over-triage, with mandatory clinician review of the most urgent categories and continuous audit of under-triage rates.

### Limitations

This study used 20 constructed referrals at a single academic centre. Although written to span the urgency spectrum, they are few relative to real volumes, cannot fully capture the heterogeneity of GP letters, and being representative rather than consecutive may not reflect real-world prevalence or ambiguity. The reference standard, from four blinded rheumatologists, embeds one service’s triage norms, which legitimately differ between countries and between services in the same jurisdiction. Model versions and prices reflect a single point in a fast-moving field, and cost was serving price rather than a health-economic evaluation. Finally, we ran no contemporaneous head-to-head human arm, so the comparison with human triage rests on external benchmarks from varied settings and taxonomies [6–10].

### Future directions

Priorities follow from these limitations: prospective evaluation on a larger consecutive sample of real de-identified referrals; external validation across services with differing triage norms; systematic optimisation of the advanced prompt, including the number and selection of worked examples and self-consistency across repeated sampling [18, 26]; formal characterisation of under-triage risk and the human-oversight workflow needed to contain it; and prospective assessment of clinician time saved and effects on waiting times. Given the pace of model development, a standing platform-based benchmarking capability, such as was used in this study, would let a service re-evaluate models and prompts as they evolve.

### Conclusion

Across 23 contemporary LLMs, the accuracy of rheumatology referral urgency triage depended strongly on both model capability and prompt design. Under a simple prompt, larger and more expensive models performed best and accuracy tracked cost. An advanced prompt substantially improved smaller and older model performance, reduced between-model variance 5.3-fold and eliminated the accuracy-cost relationship, indicating that advanced prompting can partially substitute for the inherent reasoning advantage of larger models. Set against published benchmarks, the leading models fell within or above the range reported for human referral triage, which is itself imperfect and only moderately reproducible. Triage is a substantial administrative burden that consumes rheumatologist time without advancing patient care; these data indicate that the tools required to automate it to at least a human standard already exist may well be affordable, provided under-triage is contained by appropriate validation, governance and human oversight.

## Declarations

The author has no competing interests or funding declarations.

## Data Availability

All data produced are available online at https://doi.org/10.5281/zenodo.21754374

